# Preparing for the Future: A Mixed Methods Study Protocol on AI Awareness and Educational Integration in Qatar’s Primary Health Care Workforce

**DOI:** 10.64898/2026.03.06.26347773

**Authors:** Muslim Abbas Syed, Ahmed Sameer Alnuaimi, Dana Bilal El Kaissi, Mohamed Ahmed Syed

## Abstract

**Background:** Artificial intelligence (AI) is increasingly being integrated into healthcare systems, with growing applications in clinical decision support, workflow optimization, and population health management. While substantial investments have been made in digital infrastructure, the successful adoption of AI in primary care depends critically on the readiness, awareness, and educational preparedness of healthcare professionals. Global health authorities emphasize the need for ethically grounded and workforce-focused approaches to AI integration; however, evidence on clinicians’ readiness for AI, particularly in primary care settings and in the Middle East region, remains limited.

**Objectives:** This study aims to assess the level of awareness, perceptions, attitudes, and educational needs related to AI among healthcare professionals working within Qatar’s Primary Health Care Corporation (PHCC). In addition, it seeks to examine organizational factors influencing the integration of AI-focused education in primary care and to develop an AI readiness framework that can inform targeted training strategies and policy planning.

**Methods:** This study will adopt a mixed-methods design guided by the Organizational Readiness for Change (ORC) framework, adapted for AI integration in primary care. The quantitative component will consist of an anonymous, census-style online survey distributed to all healthcare professionals across PHCC health centers and headquarters, assessing AI awareness, attitudes, training needs, and perceived infrastructure readiness. Composite AI awareness and attitude scores will be calculated, and regression analyses will be used to explore factors associated with AI readiness. The qualitative component will include semi-structured interviews and focus group discussions using maximum variation sampling to capture diverse professional perspectives. Qualitative data will be analyzed thematically, following COREQ and SRQR reporting standards. Quantitative and qualitative findings will be integrated to generate an AI readiness profile and an actionable education roadmap aligned with national digital health priorities.

**Discussion:** This study will provide the first comprehensive assessment of AI readiness among primary care healthcare professionals in Qatar. By identifying knowledge gaps, training priorities, and organizational enablers and barriers, the findings are expected to inform the development of evidence-based AI education strategies within continuing professional development frameworks. The proposed AI readiness framework may also offer a transferable model for other health systems seeking to align workforce development with responsible AI implementation in primary care.

## Introduction

Globally Artificial Intelligence (AI) has emerged as an innovative technology with wider applications in healthcare settings ranging from diagnostics and treatment planning to administrative efficiency. As AI tools become increasingly utilized in clinical workflows, the imperative for healthcare professionals to understand, accept, and effectively utilize these technologies is advocated by global health authorities such as the World Health Organization (WHO), which suggest ethically grounded, capacity-building approaches to AI integration[1, 2]. Moreover, the urgent need for structured AI education within medical curricula to ensure safe, equitable, and competent adoption across healthcare systems is increasingly emphasized in recent published literature.[3, 4]

The literature reports varied perceptions among healthcare professionals (HCPs) regarding the integration of AI. A comprehensive review documented clinicians were optimistic and favored AI applications that assist with data interpretation and workflow efficiency. HCPs is a broad category that mainly includes physicians, nurses, dentists, pharmacists, allied health care professionals, support staff that assist in delivering care and public health experts[5]. However, they expressed concerns over AI tools that directly influence clinical decisions or compromise patient relationships, privacy breaches, liability for AI-related errors, and the erosion of clinician autonomy.[6]

Importantly, studies are being initiated in the state of Qatar exploring the awareness and attitudes of healthcare professionals toward AI. For example, a recent cross-sectional survey conducted at the Primary Health Care Corporation (PHCC) reported that 78.4% of participating family physicians were aware of AI applications, with most recognizing its potential to enhance operational efficiency and healthcare delivery[7]. However, the study also highlighted gaps in formal education and training, suggesting that awareness does not necessarily equate to readiness or competence in AI use.

Qatar’s healthcare infrastructure is undergoing a digital transformation aligned with the Qatar National Vision 2030. The country has implemented a centralized electronic health record (EHR) system covering most of its prestigious healthcare institutions, enabling real-time data access and facilitating AI integration. While infrastructure readiness is advancing, the successful integration of AI into primary care hinges on targeted education, stakeholder engagement, and strategic planning. Therefore, this study will significantly contribute towards evidence base by using a mixed methods approach to comprehensively assess awareness, perceptions, and readiness, ultimately informing policy and curriculum development for AI in Qatar’s primary healthcare sector.

The primary aim of this study is to examine the level of awareness, perceptions, and educational needs related to Artificial Intelligence (AI) among healthcare professionals within Qatar’s Primary Health Care Corporation (PHCC). In addition, the study seeks to explore organizational factors that may influence the successful integration of AI-focused education into primary care practice. By assessing clinicians’ baseline knowledge and attitudes toward AI and identifying gaps in training, the research will provide evidence to inform targeted educational strategies aligned with national digital health priorities.

I. To measure the current level of awareness, understanding, and exposure to Artificial Intelligence (AI) applications among healthcare professionals working in PHCC.
II. To explore healthcare professionals’ perceptions, attitudes, and educational needs regarding the integration of AI tools in clinical practice within primary care settings.
III. To examine organizational factors within PHCC that influence the integration of AI-focused education, including perceptions of infrastructure adequacy and workforce preparedness, to identify contextual enablers and barriers to implementing effective training strategies.

### Methodology Study design

To guide the assessment of readiness, this study will apply the Organizational Readiness for Change (ORC) model, adapted for AI integration in primary care. The framework considers two interrelated dimensions: Infrastructure Readiness (technical capacity, IT systems, and resource allocation) and Human Readiness (awareness, attitudes, and competency gaps among healthcare professionals). These dimensions will be evaluated through a mixed-methods approach, combining survey data and qualitative insights. The findings will be synthesized into an AI Readiness Score, which will inform an actionable roadmap for education and phased implementation aligned with national digital health priorities. The rationale for employing mixed methods design is justified by the complexity of the research objectives, which require both numerical data to quantify awareness and readiness levels, and narrative data to explore perceptions, attitudes, and contextual factors influencing AI education integration. This design allows for triangulation of findings, enhancing the validity and depth of the study.

### Study setting

The study settings will involve all the Primary Health Care Centers (except AlJumailiya and Leghwairiya) and PHCC Headquarters AlJumailiya and Leghwairiya are excluded due to very small staff counts (<50) which would add logistical cost with little impact on representativeness relative to PHCC’s ∼7,975 employees; the exclusion is therefore pragmatic and statistically proportionate.

### Study population

All the PHCC healthcare professionals working in primary health care centers and Headquarters. The healthcare professionals will include Physicians, Dentists, Nurses, Pharmacist, Radiology technicians, Laboratory technicians, Physiotherapists, Dietitians, Health educators, administrative & support staff, and other paramedics.

### Inclusion and exclusion criteria

All health care professionals in the selected health centers including Physicians, Dentists, Nurses, Pharmacist, Radiology technicians, Laboratory technicians, Physiotherapists, Dietitians, Health educators, administrative & support staff, and other paramedics. Two PHCC health centers, AlJumailiya and Leghwairiya—will be excluded from the study because each employ fewer than 50 healthcare professionals. Including these centers would add logistical complexity without significantly contributing to the representativeness of the sample, given their very small workforce relative to the overall PHCC population of approximately 7,975 employees. Exclusion ensures efficient resource utilization while maintaining the study’s focus on centers with sufficient staff to provide meaningful insights into AI awareness and educational needs across primary care settings.

### Sampling

For the quantitative component, the study will adopt a census-style invitation. The HCPs will be approached initially through their official email with link to participate in the online survey. They will be sent a reminder in subsequent weeks. After two weeks if the response rate is low data collectors will approach HCPs with QR-code flyers which will be distributed at each center and PHCC Headquarters. In the qualitative component of this study, a maximum variation sampling strategy is employed to ensure a broad and diverse representation of healthcare professionals working in PHCC centers. This technique involves deliberately selecting participants who differ across key characteristics such as professional role (e.g., physicians, nurses, pharmacists, dentists), years of clinical experience, past exposure to AI tools, and geographic location of PHCC center.

### Sample size calculation

The current submission targets the whole population (the Healthcare Professionals working in PHCC). That is why no sample size calculation is included. We hope to receive between 1,000 and 2,000 responses.

However, the minimum sample size of 367 is known to be sufficient for calculating a prevalence estimate of 50% with 95% confidence in a small finite population of 7975 (all PHCC employees, 2024), accepting an error of 5%. Considering the important scope of this study, laying the foundation for future interventions one should consider increasing the sample size to address a wider spectrum of prevalence estimates with the same precision (error margin of 10% of the estimate magnitude).

| Sample size | Finite population size | Confidence interval (%) | Prevalence estimate (%) | Error margin (%) |
| --- | --- | --- | --- | --- |
| 367 | 7975 | 95 | 50 | 5 |
| 1080 | 7975 | 95 | 25 | 2.5 |
| 2414 | 7975 | 95 | 10 | 1 |

If the email strategy succeeds in obtaining at least 1000 responses the data collection will stop by achieving a moderate study power (table above). If, the second step of personal invitation of targeted sample is used then the study will aim to the highest study power associated with 2000 sample size (table above). This sample size will used as a stopping criterion for the study.

The formula for sample size calculation estimating a single proportion:

Finite population:

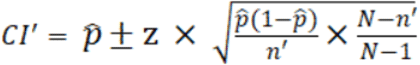

where

**z** is z score

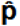is the population proportion

**n** and **n’** are sample size

**N** is the population size

For qualitative component, data will be collected till saturation of data is achieved[8]. Literature suggests that saturation is achieved normally between 12-20 interviews while conducting thematic analysis[8]. Two focus group discussions will be held with 10-15 participants in each session.

### Data collection for Quantitative online survey

An online questionnaire form will be used for data collection (supplementary document 1). The contents (the domains and items) of the online survey were constructed based on key themes and concepts discussed in recent published literature (and validated tools utilized) in this area of research.[9-13]. The questionnaire form will start with a section about demographics (age, sex, professional role, and length of clinical experience). The next section will gather information measuring awareness and knowledge about AI applications in health care. A total of 7 questions will assess the self-rated AI knowledge, exposure to AI tools and identified sources of AI knowledge.

The form includes a section on training and educational needs. A total of 4 questions in this section is dedicated to enquiring about having formal AI training, the preferred training format Workshops, Online modules, CME), identifying possible barriers for not being trained in AI. The next section of the form is exploring perceptions and attitudes. It probes the study participants about their trust in AI tools (measured on a Likert scale), the perceived benefits of AI, and AI related concerns. The remaining sections of the questionnaire form are assigned to enquire about readiness, Impact of AI on clinical decision making and patient care, and the implementation and integration of these tolls. The tool was developed by reviewing the literature.[14-18]

### Tool validation plan

The face validity is defined as the extent to which a test or scale is subjectively viewed by the targeted study participants as covering the concept it purports to measure of the designed questionnaire. Face validity answers the question “Does it appear to the test taker that the questions on the test are related to the purpose for which the test is being given?”. This aspect of the tool validation will be assessed in a PILOT study. A total of 10 prospective participants (clinicians) will be enrolled for this pilot study. Their feedback on the questionnaire will be recorded and necessary amendments made (supplementary documents 2 & 3).

The content validity is the extent to which the questions on a test are representative of the construct, trait, or attribute being measured. One way of obtaining a quantitative assessment of this type of validity is by examining the extent to which experts agree on the relevance of the content of the test items. A panel of 10 experts will be invited to review the study tool. For each questionnaire item, the panelist indicates whether the specific knowledge/ Attitude / Practice item measured is essential, useful but not essential, or not necessary as a contributor to the overall scale measure of that construct. In addition, the panelist indicates the reason for unfavorable (not essential) rating and writes any suggestion to improve the phrasing of the question. The reasons for unfavorable opinion on the side of the panelist were coded as 1=irrelevant to the construct tested, 2=too technical and difficult to the audience, 3=repeated elsewhere and 4=others (specify). The study tool will undergo iterative amendments and improvements based on the structured panelist review feedback.

The content validity ratio (CVR) for each item is computed. To determine whether an item is essential, its minimum value is compared with minimum values that depend on the number of experts who contributed ratings (a value of 0.62 for a panel of 10 experts)[[19]]. Changes required to the questionnaire will be submitted to the IRB for “Amendment Review”.

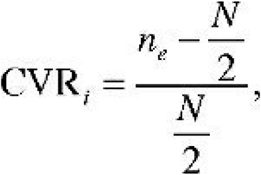

Where:

CVR_i_ = value for an item on the test

n_e_ = number of experts indicating that an item is essential

N = total number of experts in the panel

### Data collection for qualitative component of the study (interviews and FGD with healthcare professionals)

#### i Interviews with healthcare professionals

The interviews will be conducted by the PI, who have previous experience in conducting personal interviews and FGD. This process will comply with the COREQ and SRQR standards required by the researcher conducting qualitative research. The interviews are semi-structured and will be conducted utilising an interview guide and script (supplementary document 4). The interviews will last between 45 to 60 minutes.

#### ii FGD with healthcare professionals

The FGD will follow a structured discussion delineated in the FGD guide (supplementary document 5). The themes covered include assessing AI awareness and knowledge, examining training and educational needs, exploring perceptions and attitudes towards ai in healthcare, evaluating the readiness of primary care infrastructure, determining the impact of AI education on clinical decision-making and patient care, and proposing recommendations for enhancing AI education in primary care.

### Quantitative data analysis plan (online survey utilizing Microsoft Forms)

The following variables will be extracted and analyzed from the survey responses:

#### Demographics

Age, Sex, Profession (Physician, Dentist, Nurse, Pharmacist, Other)

#### AI Awareness and Knowledge

- Q7-Self-rated AI knowledge (Ordinal 4 points scale=Not familiar to Very familiar)
- Q9-AI literacy and exposure to AI tools (a list of 13 AI in healthcare application categories)
- Q8-Sources of AI knowledge (a maximum of 4 preset choices)

##### Training and Educational Needs

- Q11-Adequacy of formal AI training in practice (ordinal scale)
- Q12-Preferred training format for a healthcare provider (select between 3 choices: hands on workshops, online courses, case-based learning)
- Q13-The type of core AI competencies required from healthcare professionals in primary care: foundational knowledge, critical appraisal, medical decision making, technical use, patient communication, awareness of unintended consequences.
- Q14-Preferred structure/method of AI training delivery (Introductory AI modules integrated into medical curricula, AI-enhanced simulation training at undergraduate and postgraduate level, CPD Courses: Fundamentals of AI in Healthcare, Evidence-based CPD relating to AI in healthcare aligned with PHCC’s strategic goals and national standards, AI-focused workshops and seminars for clinical staff)

##### Attitudes/Perceptions

- Q10-Trust in AI tools (5 grades Likert scale: Strongly disagree to strongly agree)
- Q13-Unmet need for training in AI tools. The type of core AI competencies required from healthcare professionals in primary care: foundational knowledge, critical appraisal, medical decision making, technical use, patient communication, awareness of unintended consequences.
- Q15-Level of AI skills required for a healthcare practitioner (basic, proficient, expert).
- Q17-Perceived benefits/impact of AI tools on clinical practice in primary care (Improve diagnostic accuracy, Reduce time spent on documentation, Enhance patient risk stratification, Will improve patient centered care)
- Q18-Reported concerns about AI use (Risk of bias or errors in AI recommendations, Lack of transparency in AI algorithms, Reduced clinician autonomy, Patient trust and acceptance of AI)
- Q19, Q20, Q21, and Q22, Perceived infrastructure readiness to and barriers for implementing AI tools in primary care practice

#### Data analysis plan highlighting various components (variables) in the online survey

The detailed procedures for analyzing and interpreting the AI awareness and knowledge scores, as well as the logistic regression model predicting AI readiness among clinicians in Qatar is described below. The analysis will be conducted after completion of the online survey distributed across PHCC centers.

AI Awareness and Attitude Scores

#### Data Preparation

- Responses to AI awareness, knowledge and attitude items will be extracted from the survey dataset.
- Items will be coded numerically.
- Composite scores will be calculated by summing or averaging relevant items.
- All the scores will be adjusted to have a maximum possible score of 100 by multiplying the resulting total score by 100/total actual score.
- Missing data will be handled using listwise deletion.
- The AI awareness composite score is calculated by summing the scores on the 13 listed items in Q9 (a score of zero is given for “don’t know about it”, 1 point for “being aware about it” and 2 points for “having used it”.
- The AI attitude composite score, which is meant to illustrate a positive attitude with increasing score value is calculated by summing the scores on the following questions:
  1. Q10-Trust in AI tools. The first two items, AI can improve clinical outcomes, and AI tools are trustworthy are given 1 point for “Strongly Disagree” up to 5 points for “Strongly Agree” to reflect positive attitude. The third item, I’m concerned about the use of AI in patient care is inversely graded and given 1 point for “Strongly Agree” up to 5 points for “Strongly Disagree” to align with the score measuring positive attitude.
  2. Q13-Unmet need for training in AI tools. Each of the 6 core competencies are given 1 point for “Strongly Disagree” up to 5 points for “Strongly Agree”.
  3. Q15-Level of AI skills required for a healthcare practitioner (0 points for basic, 5 points for proficient, and 10 points for expert). This scoring scheme is chosen to align with the LIKERT scale maximum score of 5 per item.
  4. Q17-Perceived benefits/impact of AI tools on clinical practice in primary care. Each of the four items is given 1 point for “Strongly Disagree” up to 5 points for “Strongly Agree” to reflect positive attitude.
  5. Q18-Reported concerns about AI use. Each of the four items is inversely graded and given 1 point for “Strongly Agree” up to 5 points for “Strongly Disagree” to align with the composite score measuring positive attitude
  6. Q20-How likely are you to use AI tools if they are integrated into your clinical workflow, is given 1 point for “Very Unlikely” up to 5 points for “Very Likely” to reflect positive attitude.
  7. Q22-Perceived barriers for implementing AI tools in primary care practice. Each of the 4 items is graded inversely and given 1 point for “Strongly Agree” up to 5 points for “Strongly Disagree” to align with the composite score measuring positive attitude.

#### Statistical Methods

Descriptive statistics: for quantitative data that is assumed to be normally distributed the mean, and standard error is calculated. The median and interquartile range will be used to describe the calculated composite index scores, which will depart from the assumption of normality. The Smirnov-Kolmogorov test for alignment of a distribution with normal assumption will be used.

Group comparisons: Independent t-tests or ANOVA to compare scores across demographic groups (e.g., profession, experience) for outcome variables that are assumed to be normally distributed. The non-parametric tests (Mann-Whitney U or Kruskal-Wallis) will be used if normality assumptions are violated.

#### Qualitative data analysis

The qualitative data collected from the Focus Group Discussions and interviews will be analyzed utilizing thematic analysis.

The coding framework is derived from the themes outlined in the FGD and interview guides. Initial codes will be developed based on the following thematic areas:

1. Awareness and Knowledge of AI
2. Training and Educational Needs
3. Perceptions and Attitudes toward AI
4. Infrastructure and Readiness
5. Impact on Clinical Decision-Making and Patient Care
6. Recommendations for Enhancing AI Education

The qualitative data once recorded will be transcribed verbatim, and then analyzed using Braun and Clarke’s (2006) Thematic Analysis. This approach encompasses (1) Familiarization with data; (2) Generating initial codes; (3) Searching for themes; (4) Reviewing themes; (5) Defining and naming themes; (6) Producing the report. Two researchers will independently code an initial subset (≈20%) and discuss discrepancies; a refined codebook will then be applied to the rest.

The study will report the results in accordance with ‘Consolidation criteria for reporting qualitative research (COREQ) and ‘Standards for reporting qualitative research ‘(SRQR) guidelines.[20, 21]

Quantitative results will identify patterns and gaps; interviews/FGDs will explain the “why” and co-prioritize training content/delivery; integration products will include joint displays mapping survey findings to qualitative themes and the resulting education roadmap.

#### Quality control measures and good practices

The REPORT statement, which is an extension of the STROB statement checklist (international, collaborative initiative of epidemiologists, methodologists, statisticians, researchers and journal editors involved in the conduct and dissemination of observational studies, with the common aim of Strengthening the Reporting of Observational studies in Epidemiology) specially designed to assure the quality of reporting of secondary data analysis will be followed during analysis and writing of the report. For the qualitative component we aim to report the results in accordance with ‘Consolidation criteria for reporting qualitative research (COREQ) and ‘Standards for reporting qualitative research ‘(SRQR) guidelines.

To ensure the ethical and regulatory integrity of the study, oversight mechanisms will be implemented throughout its duration. A designated research monitor from the Clinical Research Department will conduct periodic monitoring and auditing activities. These will verify that the study is being conducted in accordance with the protocol approved by the Institutional Review Board (IRB) and outlined by Ministry of Public Health (MoPH) Qatar.

#### Timelines for study

The study is tentatively expected to be initiated in April 2026. The tentative timelines for the main study tasks are summarized in the table below:

| Components of the research study | Estimated completion dates (tentative) |
| --- | --- |
| Participant recruitment | April 2026- May 2026 |
| Data collection | May 2026- July 2026 |
| Drafting of results | August 2026-Nov 2026 |

#### Ethical Considerations

This study will be conducted in accordance with established ethical principles to ensure voluntary participation, minimize potential harm, and protect participant confidentiality. The principal investigator (PI), who will conduct the in-depth interviews and facilitate the focus group discussions (FGDs), will not hold any supervisory, managerial, or evaluative role over the recruited healthcare professionals, thereby avoiding any power relationship that could influence participation or responses. Participation in the online survey will be anonymous, and the provision of any contact information for follow-up communication will be entirely voluntary and at the discretion of the participant. The online questionnaire will explicitly state that participation is voluntary, that responses will not be linked to employment records, and that participation does not constitute an official Primary Health Care Corporation (PHCC) work task.

Specific measures will be implemented to protect potentially vulnerable participants, particularly given the workplace-based nature of the study. Recruitment will be conducted through general announcements and institutional email communications, and line managers will not be involved in recruiting their staff. Participation will be non-evaluative and will have no impact on employment status, professional standing, or performance appraisal. FGDs will be scheduled without the presence of supervisors or managers, and participants will be informed of their right to decline to answer any question and to withdraw from the study at any time without penalty or adverse consequences. All data will be deidentified prior to analysis, and study findings will be reported only in aggregate form to prevent individual identification.

Data confidentiality and security will be maintained through strict data management procedures. Survey data, audio recordings, and interview transcripts will be stored on PHCC’s secure network drive and will be accessible only to authorized members of the study team through role-based permissions. No study data will be transmitted via email or shared outside PHCC systems. A fully de-identified dataset will be used for all analyses and reporting. Study data will be retained for a period of three years following study completion, after which it will be securely deleted in accordance with PHCC data retention and disposal policies.

Ethics approval was received to conduct the study from Institutional Review Board of Primary Health Care Corporation (supplementary document).

#### Dissemination plan of study findings

Upon completion of the study, findings will be disseminated through multiple channels to ensure broad and meaningful impact. Results will be submitted for publication in peer-reviewed journals and presented at relevant scientific conferences. In addition, tailored summaries will be shared with key stakeholders, including healthcare providers and policymakers, to inform practice and decision-making. Where appropriate, public-facing materials will be developed to communicate findings to the public, thereby enhancing transparency and promoting community engagement with the research outcomes.

#### Patient and Public Involvement

There was no Patient and Public Involvement (PPI) while formulating the study tools and designing the research methodology as the study mainly focuses on AI readiness among healthcare professionals providing primary care within the state of Qatar. However, a total of 10 prospective participants (clinicians) will be enrolled for content and face validity of the tool utilized in the study. Their feedback on the questionnaire will be recorded and necessary amendments made (supplementary document).

## Discussion

The integration of Artificial Intelligence (AI) into healthcare is rapidly transforming clinical practice, operational workflows, and patient outcomes worldwide. In primary care, AI tools offer promising applications from diagnostic support and predictive analytics to administrative efficiency and personalized patient engagement. However, the successful adoption of these technologies is dependent not only on infrastructure but critically on the awareness, readiness, and education of healthcare professionals.

In Qatar, the Primary Health Care Corporation (PHCC) serves as the key organization for providing state of the art primary care services, employing a diverse workforce of healthcare professionals across 31 health centers. To the best of our knowledge there is limited evidence of how prepared this workforce is to engage with AI technologies. There is a need for a comprehensive assessment of the knowledge, perceptions, and educational needs of PHCC clinicians regarding AI and evaluation of the readiness of primary care infrastructure to support AI integration.

This study addresses that gap by employing mixed methods approach by surveying all PHCC healthcare professionals (physicians, dentists, nurses, allied health, etc., constructing and validating an AI Readiness Score to quantify readiness as primary outcome, using focus groups and interviews to identify contextual barriers/facilitators to AI education and implementation and producing an actionable AI-education roadmap aligned with national digital-health priorities. This comprehensive approach provides policy-ready evidence for PHCC leadership and educators to target training, sequence implementation, and monitor readiness over time. The study aims to explore healthcare professionals’ current understanding of AI, their attitudes toward its use in healthcare, and the systemic barriers to AI education and implementation. The inclusion of all healthcare professionals within PHCC ensures a holistic view across disciplines and roles.

Qatar’s Digital Health Strategy and National Health Strategy 3 (NHS3) place strong emphasis on digital transformation and the use of AI to improve healthcare delivery. These national priorities call for more than just advanced technology—they require a workforce that is ready and able to use these tools effectively. As the largest provider of primary care in Qatar, PHCC is central to achieving these goals. However, while infrastructure such as the centralized Electronic Health Record (EHR) system is in place, there is little evidence about how prepared PHCC’s clinicians are to adopt AI in their daily practice.

This gap matters because technology alone cannot drive change. If healthcare professionals lack the knowledge, confidence, or training to use AI tools, adoption will be slow and may even face resistance. To address this, our study will look at both technical readiness and human readiness—ensuring that investments in infrastructure are matched by efforts to build skills and confidence among clinicians.

To guide this work, we will utilize the Organizational Readiness for Change (ORC) model[22, 23], which helps assess readiness across two key areas:

- Infrastructure Readiness: Do we have the systems, IT support, and resources needed for AI integration?
- Human Readiness: Are clinicians aware of AI, do they trust its benefits, and what training do they need to feel competent?

By applying this framework, we will create an AI Readiness Score that combines these factors, identify barriers and enablers, and develop a practical roadmap for education and phased implementation. This approach ensures that PHCC’s workforce development aligns with national policy and supports safe, effective use of AI in primary care (figure 1).

**Figure 1.**
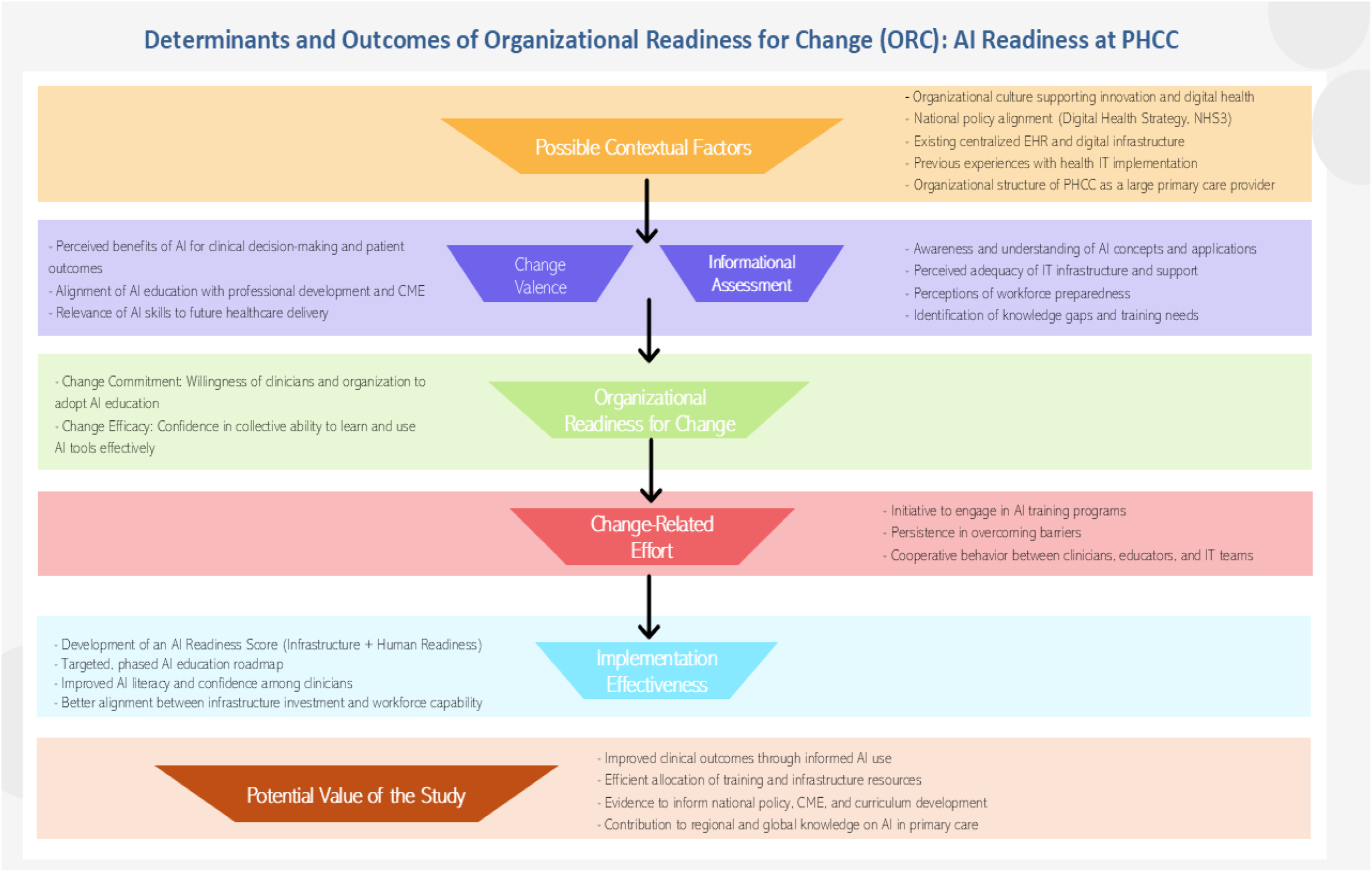
ORC framework for AI readiness at PHCC.

Enhancing clinicians’ literacy in artificial intelligence has the potential to positively influence clinical outcomes by enabling more informed and effective use of digital tools in decision-making and patient care. As AI technologies become increasingly integrated into primary care settings, clinicians’ ability to critically understand, interpret, and appropriately apply these tools is essential to ensuring their safe and effective use. By strengthening AI literacy, healthcare professionals may be better positioned to augment clinical judgment rather than rely uncritically on automated systems, thereby supporting quality of care and patient safety. In this context, assessing clinicians’ readiness for AI adoption provides valuable insight into current competencies and gaps that may influence the clinical impact of emerging technologies (figure 1).

In addition, the findings of this study have important implications for health system planning and policy development. Understanding variations in readiness and perceived competence can support more efficient allocation of resources by guiding targeted investments in training, digital infrastructure, and organizational support. At a policy level, the results may inform national strategies aimed at integrating AI education into continuing medical education and professional development frameworks, ensuring that workforce upskilling keeps pace with technological change. As one of the first empirical investigations of AI readiness among primary care clinicians in the region, this study also contributes to the global literature on AI in healthcare, offering context-specific evidence that may be informative for other health systems facing similar challenges in AI implementation.

## Data Availability

No datasets were generated or analysed during the current study. All relevant data from this study will be made available upon study completion.

